# Comparative Outcomes of Laparoscopic Fundoplication and Magnetic Sphincter Augmentation for GERD: A Systematic Review and Meta-Analysis

**DOI:** 10.1101/2025.11.10.25337526

**Authors:** Iria Cassia Abreu da Costa, Marcos Antonio Dias Vilela, Nathan Joseph Silva Godinho, Anderson Aurélio Fioreze, Sjaak Pouwels

## Abstract

**Introduction:** Patients with gastroesophageal reflux disease (GERD) who fail conservative therapy often undergo surgical intervention. While laparoscopic fundoplication (LF) is considered the standard procedure, the Magnetic Sphincter Augmentation (MSA) system has emerged as a promising alternative. Debate remains regarding their comparative effectiveness and safety.

**Methods:** This systematic review and meta-analysis examined studies comparing MSA to LF for GERD. We searched PubMed, Cochrane Central, and Embase databases using a comprehensive search strategy with multiple combinations of Medical Subject Headings (MeSH) terms and keywords related to gastroesophageal reflux disease, magnetic sphincter augmentation, LINX device, and fundoplication procedures. Key endpoints included symptom relief, quality of life, complications, recurrence, reoperation, and patient satisfaction. Studies were included if they were published in English, compared MSA (LINX Reflux Management System) with LF in patients diagnosed with GERD, and reported comparative outcomes. We excluded animal studies, case reports, case series, studies without relevant comparative data, and those with overlapping populations. Two independent reviewers screened studies using Rayyan software, with discrepancies resolved by a third reviewer. Data extraction was performed systematically using a standardized form, and quality assessment was conducted using the ROBINS-I tool for non-randomized studies. Meta-analysis was performed using random-effects models with risk ratios for dichotomous outcomes and standardized mean differences for continuous outcomes.

**Results:** Twelve studies, involving 11,690 patients (2,009 MSA; 9,681 LF), met inclusion criteria. MSA demonstrated significantly shorter operative times and hospital stays. Rates of postoperative complications, dysphagia, patient satisfaction, and reoperation did not differ significantly between groups. While both treatments provided comparable symptom control, MSA patients reported fewer gas and bloating issues, better ability to belch, and maintained the ability to vomit. There was no clear superiority of MSA over LF in terms of completely discontinuing proton pump inhibitors, although a slight trend favored MSA.

**Conclusion:** MSA and LF offer broadly similar outcomes for GERD, with no significant differences in safety, reoperation rates, and overall patient satisfaction. However, MSA may confer advantages in reducing operative times, hospital stays, gas-related symptoms, and preserving physiological functions such as belching and vomiting. These findings support the role of MSA as a viable alternative to LF, but further randomized controlled trials are needed to strengthen the evidence base and assist clinicians in tailored decision-making.

## INTRODUCTION

Gastroesophageal reflux disease (GERD) is characterized by the reflux of gastric contents into the esophagus, causing symptoms such as heartburn and regurgitation, and potentially leading to complications like esophagitis and Barrett’s esophagus. While lifestyle modifications and proton pump inhibitors (PPIs) represent initial management strategies, nearly thirty-five percent of patients fail to achieve adequate symptom control with these approaches, prompting consideration of surgical intervention. Currently, laparoscopic fundoplication (LF) remains the recommended standard surgical approach for medically refractory GERD.

The LINX Reflux Management System, a Magnetic Sphincter Augmentation (MSA) device comprising a ring of magnetic beads placed around the lower esophageal sphincter (LES), has emerged as a promising alternative. MSA is designed to augment LES pressure while preserving the ability to belch and vomit, potentially offering physiological advantages. Despite growing interest, a definitive consensus regarding its superiority over LF is lacking, in part due to heterogeneity in study designs and outcome reporting.

This systematic review and meta-analysis aimed to provide a comprehensive comparative assessment of MSA and LF. By examining outcomes including symptom relief, quality of life, complications, reoperation, and patient satisfaction, the present analysis seeks to assist clinicians and surgeons in making evidence-based decisions regarding the optimal surgical approach for GERD.

## METHODS

### Protocol Registration and Reporting Guidelines

This systematic review and meta-analysis was conducted in accordance with the Preferred Reporting Items for Systematic Reviews and Meta-Analyses (PRISMA) guidelines. The protocol was not prospectively registered.

### Search Strategy

A comprehensive systematic search of three electronic databases was conducted: PubMed (MEDLINE), Cochrane Central Register of Controlled Trials (CENTRAL), and Embase. The search was designed using the PICOT framework to ensure systematic coverage: Population consisted of patients with gastroesophageal reflux disease, Intervention was magnetic sphincter augmentation using the LINX Reflux Management System, Comparison was laparoscopic fundoplication including Nissen and Toupet variants, Outcomes included complications, operative time, length of hospital stay, postoperative proton pump inhibitor usage, reoperation rates, and patient satisfaction, and Types of studies included comparative studies while excluding case reports, case series, and animal studies. No date restrictions were applied to the search. The final search was conducted in December 2024.

The complete search strategy was as follows: (“Gastroesophageal Reflux” OR “GERD” OR “chronic GERD” OR “acid reflux” OR “reflux esophagitis” OR “peptic esophagitis” OR “esophageal reflux” OR “Esophageal Reflux” OR “Gastric Acid Reflux” OR (“Acid Reflux” AND “Gastric”) OR (“Reflux” AND “Gastric Acid”) OR “Gastric Acid Reflux Disease” OR “Gastro-Esophageal Reflux” OR “Gastro Esophageal Reflux” OR (“Reflux” AND “Gastro-Esophageal”) OR “Gastro-oesophageal Reflux” OR “Gastro oesophageal Reflux” OR (“Reflux” AND “Gastro-oesophageal”) OR “Gastroesophageal Reflux Disease” OR “GERD” OR (“Reflux” AND “Gastroesophageal”) OR “Gastro-Esophageal Reflux Disease” OR “Gastro Esophageal Reflux Disease” OR “Gastro-Esophageal Reflux Diseases” OR (“Reflux Disease” AND “Gastro-Esophageal”) OR “Heartburn” OR “Fundoplication” OR “Esophageal Sphincter, Lower” OR “Laryngopharyngeal Reflux” OR “Respiratory Aspiration of Gastric Contents” OR “Non-Erosive Reflux Disease”)

AND

(“LINX Reflux Management System” OR “LINX” OR “magnetic sphincter augmentation” OR “titanium beads” OR “magnetic lower esophageal sphincter augmentation” OR “esophageal sphincter device” OR “magnetic LES augmentation” OR “LES device” OR “MSA” OR “magnetic sphincter” OR “LINX device” OR (“magnetic augmentation” AND “sphincter”) OR (“magnetic augmentation” AND “LES”) OR (“magnetic augmentation” AND “lower esophageal sphincter”) OR “LINX system” OR (“magnetic device” AND “GERD”) OR (“magnetic device” AND “reflux”))

AND

(“Fundoplication” OR “fundoplication” OR “Nissen fundoplication” OR “Toupet fundoplication” OR “video-assisted fundoplication” OR “laparoscopic fundoplication” OR “surgical reflux management” OR “anti-reflux surgery” OR “partial fundoplication” OR “complete fundoplication” OR “laparoscopic anti-reflux surgery” OR “LARS” OR “Nissen Operation” OR “Operation, Nissen” OR (“fundic wrapping” AND “Gastroesophageal Reflux”) OR (“fundic wrapping” AND “hiatal hernia”) OR “lower esophagus mobilization” OR “esophageal mobilization” OR “esophageal plication” OR “fundic plication”).

The search strategy was developed iteratively with input from a medical librarian and was peer-reviewed by all authors. Search terms were tested for sensitivity by ensuring that key landmark studies in the field were captured. No language restrictions were initially applied during the search phase, but only English-language articles were ultimately included in the review due to resource constraints for translation.

### Inclusion and Exclusion Criteria

Studies were considered eligible for inclusion if they met all of the following criteria: the study population consisted of adult patients (age eighteen years or older) with a confirmed diagnosis of gastroesophageal reflux disease based on clinical symptoms, endoscopic findings, pH monitoring, or manometry; the intervention group received magnetic sphincter augmentation using the LINX Reflux Management System as the primary surgical treatment for GERD; the comparison group received laparoscopic fundoplication including Nissen fundoplication, Toupet fundoplication, or other laparoscopic antireflux procedures; the study reported at least one comparative outcome measure of interest including but not limited to complications (intraoperative or postoperative), operative time, length of hospital stay, postoperative symptoms (dysphagia, gas bloat, ability to belch, ability to vomit), quality of life measures (GERD-HRQL scores), postoperative proton pump inhibitor usage, reoperation rates, or patient satisfaction scores; the study design was a randomized controlled trial, prospective cohort study, retrospective cohort study, or case-control study with a comparative design; and the study was published in the English language in a peer-reviewed journal.

Studies were excluded if they met any of the following criteria: animal or laboratory studies investigating magnetic sphincter augmentation or fundoplication procedures; case reports describing individual patients or case series with fewer than ten patients and no comparison group; studies that reported only the MSA group or only the fundoplication group without providing comparative data between the two interventions; studies in which the study populations overlapped substantially with another included study, in which case the study with the larger sample size or more comprehensive outcome reporting was selected; studies published in languages other than English; conference abstracts, letters to the editor, editorials, or review articles that did not present original comparative data; studies that included pediatric populations (under eighteen years of age) exclusively, although studies with mixed adult and pediatric populations were evaluated individually for inclusion; studies in which patients underwent concurrent procedures (such as hiatal hernia repair exceeding three centimeters, bariatric surgery, or other major gastrointestinal procedures) that would substantially confound the comparison between MSA and fundoplication; and studies in which outcome measures were not clearly defined or could not be extracted for meta-analysis.

### Study Selection Process

The study selection process followed a systematic two-stage screening approach. In the first stage, two independent reviewers (ICAC and MADV) screened all titles and abstracts identified through the database searches using Rayyan software, a web-based application designed for systematic review screening. Rayyan’s duplicate detection feature was used to identify and remove duplicate records. Each reviewer independently marked studies as include, exclude, or maybe based on the predetermined inclusion and exclusion criteria. The reviewers were not blinded to study authors, institutions, or journal names during the screening process. Any disagreements between the two reviewers were flagged by the Rayyan system for resolution. In cases of disagreement, a third independent reviewer (NJSG) was consulted, and the final decision was reached through consensus discussion among all three reviewers.

In the second stage, full-text articles of all potentially eligible studies identified during title and abstract screening were retrieved for detailed evaluation. Two reviewers (ICAC and MADV) independently assessed each full-text article against the complete inclusion and exclusion criteria. A standardized full-text screening form was used to document the reasons for exclusion of studies that did not meet eligibility criteria. Disagreements at the full-text review stage were again resolved through discussion with the third reviewer (NJSG) until consensus was achieved. Studies that met all inclusion criteria and did not meet any exclusion criteria were included in the final qualitative synthesis, and those with sufficient quantitative data were included in the meta-analysis. The inter-rater reliability between reviewers was assessed using Cohen’s kappa statistic and was found to be substantial (kappa equals zero point seven eight), indicating good agreement between independent reviewers.

### Data Extraction

Data extraction was performed systematically using a standardized, piloted data extraction form developed specifically for this review. The data extraction form was piloted on three studies not included in the final analysis to ensure clarity and completeness of all fields. Two reviewers (MADV and NJSG) independently extracted data from each included study, and any discrepancies were resolved through discussion with a third reviewer (ICAC). The data extraction process was managed using Microsoft Excel spreadsheets with predefined fields and drop-down menus to minimize errors and ensure consistency.

For each included study, the following study characteristics were extracted: first author name and year of publication, study design categorized as randomized controlled trial, prospective cohort study, retrospective cohort study, or case-control study, geographic location and country where the study was conducted, study period indicating the dates of patient enrollment and follow-up, sample size for both the MSA group and the fundoplication group, and length of follow-up reported in months or years. Detailed patient demographic and clinical characteristics were also extracted, including age reported as mean with standard deviation or median with interquartile range for each treatment group, sex distribution reported as number and percentage of male and female patients in each group, body mass index (BMI) reported as mean with standard deviation for each group, GERD severity assessed using validated instruments such as the GERD Health-Related Quality of Life (GERD-HRQL) questionnaire with scores reported as mean and standard deviation, objective GERD measures including DeMeester score from twenty-four hour pH monitoring, percentage of time with pH less than four, and lower esophageal sphincter pressure from esophageal manometry, duration of PPI therapy prior to surgery, hiatal hernia size categorized as absent, small (less than two centimeters), moderate (two to three centimeters), or large (greater than three centimeters), and previous abdominal or esophageal surgeries if reported.

Surgical characteristics and intraoperative data extracted included specific type of fundoplication performed in the comparison group (Nissen fundoplication, Toupet fundoplication, or other variants), concurrent procedures performed such as hiatal hernia repair or cruroplasty, operative time measured in minutes and reported as mean with standard deviation or median with interquartile range, estimated blood loss if reported, and conversion to open surgery rates. Postoperative outcomes formed the core of the data extraction and included length of hospital stay measured in days and reported as mean with standard deviation or median with interquartile range, complications categorized as early (within thirty days) or late (beyond thirty days) and further classified as major (requiring intervention or prolonging hospital stay) or minor, specific complication types including bleeding, infection, perforation, pneumonia, venous thromboembolism, and device-related complications such as erosion or migration for MSA patients, postoperative dysphagia severity graded as none, mild, moderate, or severe and measured using validated dysphagia scores when available, need for endoscopic dilation for dysphagia, gas bloat symptoms and their severity, ability to belch assessed as preserved or impaired, ability to vomit when necessary assessed as preserved or impaired, postoperative symptom control measured using GERD-HRQL scores or other validated symptom questionnaires, postoperative PPI usage categorized as complete discontinuation, normal-dose usage, or high-dose usage, patient satisfaction measured using validated satisfaction scores or binary satisfied versus not satisfied responses, quality of life improvements from baseline using validated instruments, reoperation rates including the indication for reoperation such as symptom recurrence, complications, or device malformation, and long-term outcomes when available including symptom recurrence and need for alternative treatments.

When studies reported continuous outcomes as medians with ranges or interquartile ranges instead of means with standard deviations, we used the method described by Wan and colleagues to estimate the mean and standard deviation. This approach uses sample size, median, and range or interquartile range to approximate the mean and standard deviation, allowing inclusion of these studies in the meta-analysis. For dichotomous outcomes, we extracted the number of events and the total number of patients in each group. When outcomes were reported at multiple time points, we extracted data from the longest follow-up period available to capture the most comprehensive assessment of treatment effects. In cases where data were presented only in graphical form, we used digital plot digitizer software (WebPlotDigitizer version four point six) to extract numerical values from graphs and figures.

### Quality Assessment and Risk of Bias

The methodological quality and risk of bias of included non-randomized studies were assessed using the Risk Of Bias In Non-randomized Studies of Interventions (ROBINS-I) tool. This tool evaluates bias across seven domains: bias due to confounding, bias in selection of participants into the study, bias in classification of interventions, bias due to deviations from intended interventions, bias due to missing data, bias in measurement of outcomes, and bias in selection of the reported result. Two independent reviewers (ICAC and MADV) assessed each study across all seven domains, assigning a judgment of low risk, moderate risk, serious risk, critical risk, or no information for each domain. The overall risk of bias for each study was determined based on the most severe risk of bias judgment in any domain.

Specific considerations were made for common sources of bias in surgical comparative studies. For bias due to confounding, we evaluated whether important prognostic factors such as age, GERD severity, hiatal hernia size, and previous treatments were balanced between groups or adequately adjusted for in the analysis. For bias in selection of participants, we assessed whether selection into the MSA or fundoplication group could have been related to prognostic factors. For bias in classification of interventions, we determined whether the intervention status was well-defined and whether misclassification of intervention status could have occurred. For bias due to deviations from intended interventions, we evaluated whether there were systematic differences in the care provided beyond the interventions being compared. For bias due to missing data, we assessed the completeness of outcome data and whether missing data could have introduced bias. For bias in measurement of outcomes, we evaluated whether outcome assessors were blinded to intervention status and whether outcome measurement methods were comparable between groups. For bias in selection of reported results, we determined whether reported outcomes were pre-specified and whether selective reporting of results was likely.

The results of the quality assessment were used to inform the interpretation of findings and to identify potential limitations that should be considered when drawing conclusions from the meta-analysis. Studies at critical risk of bias were retained in the analysis but sensitivity analyses were planned to assess the impact of excluding these studies on the overall conclusions. Publication bias was assessed qualitatively by examining the symmetry of funnel plots for outcomes with ten or more studies, and quantitatively using Egger’s regression test when appropriate.

### Data Synthesis and Statistical Analysis

Statistical analysis was performed using Stata version eighteen (StataCorp LLC, College Station, Texas, United States). Meta-analysis was conducted using random-effects models based on the DerSimonian and Laird method, which accounts for both within-study and between-study variability. Random-effects models were selected a priori due to the anticipated heterogeneity in study designs, patient populations, surgical techniques, and outcome measurement methods across included studies.

For dichotomous outcomes such as complications, reoperation rates, and patient satisfaction, we calculated risk ratios (RR) with ninety-five percent confidence intervals (CI) for each study. When event rates were low (less than ten percent), we also calculated odds ratios to assess the robustness of findings. The pooled risk ratio was calculated using the Mantel-Haenszel method within a random-effects framework. For continuous outcomes such as operative time, length of hospital stay, and GERD-HRQL scores, we calculated standardized mean differences (SMD) expressed as Hedges’s g with ninety-five percent confidence intervals when studies used different measurement scales, or weighted mean differences (WMD) with ninety-five percent confidence intervals when all studies used the same scale. Hedges’s g was preferred over Cohen’s d because it provides a less biased estimate in studies with small sample sizes by applying a correction factor.

Statistical heterogeneity among studies was assessed using three complementary methods. First, the I-squared (I²) statistic was calculated to quantify the percentage of total variation across studies that is due to heterogeneity rather than chance, with values of zero to forty percent considered as low heterogeneity, thirty to sixty percent as moderate heterogeneity, fifty to ninety percent as substantial heterogeneity, and seventy-five to one hundred percent as considerable heterogeneity. Second, the tau-squared (_τ_²) statistic was calculated to estimate the between-study variance. Third, the Cochran Q test was performed with a p-value of less than zero point one considered statistically significant for heterogeneity due to the low power of this test. When substantial or considerable heterogeneity was detected, we explored potential sources through subgroup analyses and meta-regression when sufficient studies were available.

Planned subgroup analyses were conducted based on study design (prospective versus retrospective studies), type of fundoplication (Nissen versus Toupet), geographic region (United States versus Europe), GERD severity (mild-to-moderate versus severe), hiatal hernia size (small versus moderate-to-large), and length of follow-up (short-term defined as less than one year versus long-term defined as one year or more). Meta-regression was performed for continuous moderator variables such as mean patient age and mean BMI when at least ten studies contributed data for a given outcome. Sensitivity analyses were conducted by sequentially removing each study and recalculating the pooled estimate to assess the influence of individual studies on the overall result, and by restricting analyses to studies at low or moderate risk of bias as determined by ROBINS-I assessment.

Statistical significance was set at a two-tailed alpha level of zero point zero five for all analyses except for tests of heterogeneity where alpha was set at zero point one. Results were reported as risk ratios or standardized mean differences with corresponding ninety-five percent confidence intervals and p-values. Forest plots were generated for all meta-analyses to visually display individual study results and pooled estimates. All analyses were conducted according to intention-to-treat principles when possible, although most included studies did not explicitly report intention-to-treat analyses.

## RESULTS

### Study Selection

The initial database searches identified a total of six hundred ninety-four records: three hundred nineteen from PubMed, three hundred sixty-seven from Embase, and eight from Cochrane Central Register of Controlled Trials. After removing one hundred ninety-one duplicates using Rayyan’s automated duplicate detection feature supplemented by manual review, five hundred three unique records remained for title and abstract screening. During the first stage of screening, four hundred seventy-four records were excluded because they clearly did not meet inclusion criteria based on title and abstract review. The most common reasons for exclusion at this stage were studies that did not include both MSA and fundoplication comparison groups, studies that were not comparative in design, review articles, conference abstracts without full-text availability, and studies in languages other than English.

Twenty-nine full-text articles were retrieved and assessed for eligibility in the second stage of screening. Of these, one article could not be retrieved despite attempts to contact the authors and library services. An additional sixteen articles were excluded after full-text review for the following reasons: ten were conference abstracts without corresponding full-text publications containing sufficient methodological detail and extractable data, five were identified as duplicate publications reporting on overlapping patient populations from the same institutions during the same time periods (in these cases, the publication with the larger sample size or more comprehensive outcome reporting was retained), and one was a systematic review article that did not contribute original comparative data. No randomized controlled trials were identified; all included studies were observational in design. Ultimately, twelve studies met all inclusion criteria and were included in the final qualitative synthesis and quantitative meta-analysis. The study selection process is summarized in the PRISMA flow diagram (Figure one).

### Study Characteristics

The twelve included studies were published between two thousand fourteen and two thousand twenty-four and comprised a total of eleven thousand six hundred ninety patients, of whom two thousand nine underwent MSA and nine thousand six hundred eighty-one underwent LF. Four studies (Callahan and colleagues published in two thousand twenty-three, Bonavina and colleagues published in two thousand twenty-one, Ayazi and colleagues published in two thousand nineteen, and Riegler and colleagues published in two thousand fifteen) employed prospective designs with either prospective cohort or prospective observational methodologies. The remaining eight studies (Wisniowski and colleagues two thousand twenty-four, Asti and colleagues two thousand twenty-three, O’Neill and colleagues two thousand twenty-two, Warren and colleagues two thousand sixteen, Reynolds and colleagues two thousand sixteen, Reynolds and colleagues two thousand fifteen, Sheu and colleagues two thousand fifteen, and Louie and colleagues two thousand fourteen) used retrospective designs including retrospective cohort studies, case-control studies, and matched-pair analyses.

Most studies were conducted in the United States, with nine of the twelve studies reporting data from American institutions. Three European investigations contributed international perspectives: Asti and colleagues from Italy in two thousand twenty-three, and two multicenter European studies by Bonavina and colleagues in two thousand twenty-one and Riegler and colleagues in two thousand fifteen, both of which included patients from Austria, Germany, Italy, and the United Kingdom. Sample sizes varied considerably across studies, ranging from small single-center cohorts of twenty-four patients in the study by Sheu and colleagues to large retrospective database analyses including seven thousand eight hundred eighty-two patients in the study by Wisniowski and colleagues. The median sample size across all studies was approximately two hundred patients.

Follow-up duration varied substantially among included studies. Short-term follow-up of less than one year was reported in three studies, medium-term follow-up of one to three years was reported in six studies, and long-term follow-up of more than three years was reported in three studies. The longest follow-up period was five years, reported by O’Neill and colleagues in two thousand twenty-two. Study characteristics for all included studies are summarized in Table one.

### Patient Characteristics

Patient demographics and baseline clinical characteristics were generally well-reported across studies, although not all studies provided complete data for all variables. Patients who underwent MSA were generally younger than those who underwent fundoplication, with mean ages ranging from thirty-nine point three years to fifty-six years in the MSA groups compared to forty-three point eight years to sixty-five point eight years in the fundoplication groups. This age difference was statistically significant in several individual studies and likely reflects patient selection based on surgeon preference, patient preference, or institutional protocols.

Sex distribution was relatively balanced between groups across most studies, with the proportion of female patients ranging from approximately forty to sixty percent in both MSA and fundoplication groups. Body mass index data were reported in seven studies and showed comparable BMI between groups, with mean values typically in the overweight range (BMI twenty-five to thirty kilograms per meter squared) for both MSA and fundoplication patients. All included studies enrolled patients with established GERD diagnosis confirmed by clinical symptoms and objective testing. Typical inclusion criteria across studies required documented pathological acid exposure on twenty-four hour pH monitoring, abnormal DeMeester scores (usually greater than fourteen point seven), and either partial response or non-response to proton pump inhibitor therapy.

Baseline GERD severity as measured by GERD Health-Related Quality of Life (GERD-HRQL) questionnaire scores was reported in four studies, with baseline scores ranging from approximately nineteen to thirty-five points, indicating moderate to severe symptom burden. DeMeester scores, which quantify esophageal acid exposure, were reported in five studies with mean baseline scores ranging from one hundred fifty-three to three hundred thirty, substantially exceeding the normal threshold of fourteen point seven. Percentage of time with esophageal pH less than four, another measure of acid exposure, was reported in three studies with mean values ranging from sixty-one to seventy-eight percent, again far exceeding the normal threshold of approximately four percent.

Duration of PPI therapy prior to surgery was reported in four studies, with patients typically having received PPI treatment for two hundred forty-three to four hundred six days (approximately eight to thirteen months) before proceeding to surgical intervention. Hiatal hernia characteristics were reported variably across studies. Most studies included patients with small hiatal hernias (less than two centimeters) or excluded patients with large hiatal hernias (greater than three centimeters), as large hiatal hernias typically required more extensive repair and were considered a relative contraindication to MSA in many studies. Detailed patient characteristics for all included studies are presented in Table two.

### Risk of Bias Assessment

Using the ROBINS-I tool, all twelve observational studies were assessed to have an overall serious risk of bias. This determination was primarily driven by concerns regarding confounding and selection bias, which are inherent limitations of non-randomized study designs. In the domain of bias due to confounding, most studies were at serious risk because important prognostic factors such as age, GERD severity, and hiatal hernia size were not balanced between groups and were not adequately adjusted for in statistical analyses. The consistent finding that MSA patients were younger than fundoplication patients across multiple studies suggests that systematic differences in patient selection occurred, likely based on surgeon judgment regarding patient suitability for each procedure.

For bias in selection of participants into the study, most retrospective studies were at serious risk because the selection of patients into MSA versus fundoplication groups was based on factors that may be related to outcomes, such as patient age, surgeon preference, or institutional protocols that evolved over time as MSA became available. The prospective studies generally had moderate risk in this domain because while selection was non-random, the criteria for selecting patients into each intervention group were more clearly defined. Bias in classification of interventions was generally low across all studies because the distinction between MSA and fundoplication procedures is clear and objective, with little possibility for misclassification.

For bias due to deviations from intended interventions, most studies were at low to moderate risk, as there was no indication of systematic differences in perioperative care between MSA and fundoplication patients beyond the surgical procedures themselves. For bias due to missing data, most studies were at low risk, with complete outcome data reported for the majority of participants and no indication that missing data was related to outcomes. For bias in measurement of outcomes, studies were generally at moderate risk because while most outcomes were objectively defined (such as operative time and length of hospital stay), some subjective outcomes (such as patient satisfaction and symptom scores) may have been influenced by patients’ and providers’ knowledge of which procedure was performed. Finally, for bias in selection of reported results, studies were generally at low to moderate risk, with no strong indication of selective outcome reporting, although many studies lacked prospectively published protocols to confirm pre-specification of outcomes.

Despite the overall serious risk of bias across studies, the consistency of findings across multiple independent studies for several key outcomes (particularly operative time and hospital length of stay) provides some reassurance regarding the robustness of these findings. However, caution is warranted in interpreting results, particularly for outcomes where only a few studies contributed data or where heterogeneity was substantial. The limitations imposed by the observational nature of all included studies underscore the critical need for large-scale, well-designed randomized controlled trials to provide more definitive evidence regarding the comparative effectiveness of MSA versus fundoplication.

### Synthesis of Results

Meta-analysis was performed for fifteen distinct outcomes for which sufficient comparable data were available from multiple studies. Not all outcomes were reported in every study, and the number of studies contributing to each meta-analysis varied from three to five studies depending on the specific outcome. Effect estimates for outcomes with fewer than three studies were not pooled but are reported narratively. Heterogeneity varied considerably across different outcomes, ranging from no heterogeneity (I-squared equals zero percent) to substantial heterogeneity (I-squared exceeding seventy-five percent). Random-effects models were used for all meta-analyses regardless of heterogeneity level, as planned a priori.

For operative time, five studies contributed data (Wisniowski two thousand twenty-four, Callahan two thousand twenty-three, Bonavina two thousand twenty-one, Reynolds two thousand sixteen, and Sheu two thousand fifteen). All five studies consistently demonstrated shorter operative times for MSA compared to fundoplication. The pooled analysis using standardized mean difference expressed as Hedges’s g yielded a value of minus one point zero three (ninety-five percent confidence interval minus one point three six to minus zero point seven zero, p-value less than zero point zero zero one), indicating a large and statistically significant reduction in operative time favoring MSA. Heterogeneity was moderate (I-squared equals fifty-eight percent). The consistency across all five studies, with effect estimates ranging from moderate to large reductions in operative time for MSA, provides strong evidence that MSA procedures are completed more quickly than fundoplication procedures. This finding is clinically meaningful as shorter operative times may reduce anesthesia exposure, operating room costs, and potentially reduce complications related to prolonged surgery.

Hospital length of stay was reported in four studies (Wisniowski two thousand twenty-four, Callahan two thousand twenty-three, Reynolds two thousand sixteen, and Sheu two thousand fifteen). The pooled standardized mean difference was minus one point zero one (ninety-five percent confidence interval minus one point seven two to minus zero point three zero, p-value equals zero point zero one), demonstrating a large and statistically significant reduction in hospital stay duration for MSA compared to fundoplication. Heterogeneity was substantial (I-squared equals eighty-four percent), which may reflect differences in institutional discharge protocols, geographic practice patterns, and insurance systems (particularly between United States and European studies). Despite this heterogeneity, the direction of effect was consistent across all four studies, with MSA patients experiencing shorter hospital stays. From a health system perspective, shorter hospital stays translate to reduced healthcare costs and earlier return of patients to their normal activities.

Overall complication rates were assessed in four studies (Wisniowski two thousand twenty-four, Bonavina two thousand twenty-one, Warren two thousand sixteen, and Reynolds two thousand fifteen). The pooled log odds ratio was minus zero point eight five (ninety-five percent confidence interval minus two point zero two to zero point three three, p-value equals zero point one six), indicating no statistically significant difference in overall complication rates between MSA and fundoplication. Individual study results were mixed, with one study (Bonavina and colleagues) showing lower complication rates with MSA, while the other three studies showed no significant difference. Heterogeneity was moderate (I-squared equals forty-three percent). This finding suggests that both procedures have comparable safety profiles regarding overall complications, although the wide confidence interval indicates substantial uncertainty and the possibility that clinically important differences may exist but were not detected with the available data.

Postoperative dysphagia was examined in four studies (Asti two thousand twenty-three, Warren two thousand sixteen, Reynolds two thousand sixteen, and Reynolds two thousand fifteen), with a pooled log odds ratio of zero point three six (ninety-five percent confidence interval minus zero point three six to one point zero nine, p-value equals zero point three three), showing no statistically significant difference between groups. Individual studies showed mixed results, with one study favoring fundoplication and three showing no difference. Dysphagia is a recognized complication of both procedures, particularly in the early postoperative period, and the similar rates between MSA and fundoplication suggest that neither procedure has a clear advantage in preventing this common side effect. However, it is important to note that dysphagia was assessed using different methods and time points across studies, which may have contributed to the heterogeneity (I-squared equals forty-nine percent).

Severe dysphagia, defined variably across studies but generally requiring medical intervention or significantly impacting quality of life, was reported in three studies (Warren two thousand sixteen, Reynolds two thousand sixteen, and Reynolds two thousand fifteen). The pooled log odds ratio was minus zero point eight three (ninety-five percent confidence interval minus one point eight nine to zero point two three, p-value equals zero point one three), showing a trend toward lower rates of severe dysphagia with MSA that did not reach statistical significance. This outcome is particularly relevant clinically because while mild dysphagia is common and often transient after antireflux surgery, severe persistent dysphagia can significantly impair quality of life and may require additional interventions such as endoscopic dilation or even reoperation.

Dysphagia requiring endoscopic dilation was reported in five studies (O’Neill two thousand twenty-two, Reynolds two thousand sixteen, Reynolds two thousand fifteen, Louie two thousand fourteen, and Sheu two thousand fifteen), with a pooled log odds ratio of zero point three six (ninety-five percent confidence interval minus zero point four two to one point one four, p-value equals zero point three six). Individual study results showed one study favoring fundoplication and four showing no difference. The need for dilation represents a more objective measure of clinically significant dysphagia compared to subjective symptom reporting, and the similar rates between MSA and fundoplication provide reassurance that neither procedure substantially increases the risk of severe dysphagia requiring intervention.

Gas bloat symptoms were evaluated using two separate measures. General gas or bloating symptoms were reported in three studies (Reynolds two thousand sixteen, Riegler two thousand fifteen, and Louie two thousand fourteen), with a pooled log odds ratio of minus one point three three (ninety-five percent confidence interval minus one point eight eight to minus zero point seven eight, p-value less than zero point zero zero one), demonstrating a statistically significant reduction in gas and bloating symptoms with MSA compared to fundoplication. Two of the three studies showed statistically significant benefits favoring MSA. This finding is clinically important because gas-bloat syndrome, characterized by abdominal bloating, inability to belch, and increased flatulence, is a well-recognized complication of fundoplication that can significantly impair quality of life. The ability of MSA to reduce these symptoms while still providing reflux control represents a meaningful advantage.

Postoperative gas bloat specifically (as opposed to general gas or bloating symptoms) was reported in three studies (Warren two thousand sixteen, Reynolds two thousand sixteen, and Reynolds two thousand fifteen) with mixed results. One study favored MSA, one favored fundoplication, and one showed no difference. The pooled log odds ratio was zero point one eight (ninety-five percent confidence interval minus one point one seven to one point five four, p-value equals zero point seven nine), indicating no statistically significant difference. Severe gas bloat was reported in three studies (Asti two thousand twenty-three, Warren two thousand sixteen, and Reynolds two thousand fifteen) with a pooled log odds ratio of zero point three one (ninety-five percent confidence interval minus zero point four four to one point zero six, p-value equals zero point four two), also showing no significant difference. The discrepancy between the findings for general gas bloating symptoms (which favored MSA) and the specific gas bloat and severe gas bloat outcomes (which showed no difference) may reflect differences in how these outcomes were defined and measured across studies.

The ability to belch when necessary was assessed in three studies (Reynolds two thousand sixteen, Riegler two thousand fifteen, and Louie two thousand fourteen), reported as inability to belch. The pooled log odds ratio was minus one point eight eight (ninety-five percent confidence interval minus two point seven four to minus one point zero two, p-value less than zero point zero zero one), demonstrating a large and highly statistically significant reduction in inability to belch with MSA compared to fundoplication. All three studies consistently showed this benefit. This finding is particularly relevant because the inability to belch is a common complaint after fundoplication and contributes to the gas-bloat syndrome. The preservation of the ability to belch with MSA represents a physiological advantage that may translate to improved patient satisfaction and quality of life.

Similarly, the ability to vomit when necessary was examined in three studies (Reynolds two thousand sixteen, Reynolds two thousand fifteen, and Riegler two thousand fifteen), reported as inability to vomit. The pooled log odds ratio was minus two point two six (ninety-five percent confidence interval minus two point nine zero to minus one point six two, p-value less than zero point zero zero one), showing a very large and highly statistically significant reduction in inability to vomit with MSA. All three studies uniformly demonstrated this advantage. While the inability to vomit may not seem as clinically significant as other outcomes, it can be important in specific situations such as food poisoning, gastroenteritis, or excessive alcohol consumption, and some patients express concern about losing this protective reflex. The preservation of the ability to vomit with MSA provides patients with greater physiological normalcy.

Discontinuation of proton pump inhibitors was reported in five studies (O’Neill two thousand twenty-two, Reynolds two thousand sixteen, Reynolds two thousand fifteen, Riegler two thousand fifteen, and Louie two thousand fourteen). Individual study results were mixed, with one study favoring MSA, one favoring fundoplication, and three showing no difference. The pooled log odds ratio was minus zero point two nine (ninety-five percent confidence interval minus one point one six to zero point five nine, p-value equals zero point five two), indicating no statistically significant difference in complete PPI cessation rates between MSA and fundoplication. Both procedures were successful in allowing many patients to discontinue PPI therapy, which is a primary goal of surgical treatment for GERD. However, the confidence interval is wide, and heterogeneity was substantial (I-squared equals sixty-seven percent), suggesting that true differences may exist but were not consistently detected across studies.

The need for normal-dose PPIs postoperatively was assessed in four studies (Asti two thousand twenty-three, O’Neill two thousand twenty-two, Warren two thousand sixteen, and Reynolds two thousand fifteen). Individually, all four studies showed no statistically significant differences between groups. However, the pooled log odds ratio of zero point four five (ninety-five percent confidence interval zero point zero three to zero point eight seven, p-value equals zero point zero four) reached statistical significance, suggesting that patients who underwent fundoplication were slightly less likely to require normal-dose PPIs postoperatively compared to those who underwent MSA. This finding should be interpreted cautiously because the difference is small, the confidence interval is narrow and only barely excludes the null value, and the clinical significance of requiring normal-dose PPI versus being completely off PPI may be debatable. Additionally, decisions about postoperative PPI use may be influenced by multiple factors including physician practice patterns, patient preferences, and symptom profiles that may not reflect true differences in procedure efficacy.

Reoperation rates were examined in four studies (Wisniowski two thousand twenty-four, O’Neill two thousand twenty-two, Warren two thousand sixteen, and Reynolds two thousand fifteen), with a pooled log odds ratio of zero point zero zero (ninety-five percent confidence interval minus one point zero six to one point zero seven, p-value equals zero point nine nine), indicating virtually identical reoperation rates between MSA and fundoplication. This finding provides important reassurance regarding the durability of MSA outcomes, as concerns about device-related complications such as erosion or migration requiring removal have been raised. The similar reoperation rates suggest that while the mechanisms of failure may differ between the two procedures (device-related issues for MSA versus wrap disruption or recurrent herniation for fundoplication), the overall likelihood of requiring reoperation is comparable.

Patient satisfaction was reported in three studies (O’Neill two thousand twenty-two, Reynolds two thousand sixteen, and Reynolds two thousand fifteen) using various satisfaction scales or binary satisfied versus not satisfied assessments. The pooled log odds ratio was minus zero point three zero (ninety-five percent confidence interval minus zero point nine seven to zero point three seven, p-value equals zero point three eight), showing no statistically significant difference in patient satisfaction between MSA and fundoplication. High satisfaction rates were reported with both procedures, typically exceeding eighty percent in both groups. This finding suggests that despite differences in specific outcomes such as gas bloating and ability to belch, overall patient satisfaction is comparable, perhaps because both procedures effectively achieve the primary goal of controlling reflux symptoms.

Willingness to undergo the procedure again was assessed in three studies (O’Neill two thousand twenty-two, Reynolds two thousand sixteen, and Reynolds two thousand fifteen) as a measure of patient satisfaction and treatment regret. The pooled log odds ratio was zero point two eight (ninety-five percent confidence interval minus zero point seven eight to one point three four, p-value equals zero point six zero), indicating no statistically significant difference between groups. The majority of patients in both groups expressed willingness to undergo their respective procedures again, with rates typically exceeding seventy percent. This measure provides a comprehensive patient-centered assessment that integrates the entire surgical experience and outcomes, and the similar rates between groups further support the conclusion that both procedures provide acceptable outcomes from the patient perspective.

GERD Health-Related Quality of Life scores were reported postoperatively in four studies (Callahan two thousand twenty-three, O’Neill two thousand twenty-two, Bonavina two thousand twenty-one, and Reynolds two thousand sixteen). The pooled standardized mean difference was zero point one eight (ninety-five percent confidence interval minus zero point two seven to zero point six two, p-value equals zero point four four), indicating no statistically significant difference in quality of life outcomes between MSA and fundoplication. Both procedures resulted in substantial improvements from baseline GERD-HRQL scores, with postoperative scores typically in the low range (less than ten points) indicating minimal residual reflux symptoms. The similar quality of life outcomes between groups suggest that both procedures are effective in relieving GERD-related symptoms and improving patient well-being.

## DISCUSSION

### Principal Findings

This systematic review and meta-analysis synthesizing data from twelve observational studies including eleven thousand six hundred ninety patients found that magnetic sphincter augmentation and laparoscopic fundoplication share broadly similar safety and efficacy profiles in treating gastroesophageal reflux disease. However, MSA provided certain procedural and symptomatic advantages, including significantly shorter operative durations, shorter hospital stays, fewer gas-related symptoms, and improved preservation of physiological functions such as belching and vomiting. Other critical outcomes, including overall postoperative complications, dysphagia, reoperation rates, patient satisfaction, and health-related quality of life, did not differ significantly between the two procedures. These findings suggest that MSA represents a viable alternative to traditional fundoplication, particularly for appropriately selected patients who prioritize faster recovery and preservation of normal physiological functions.

### Comparison with Existing Literature

The findings of this meta-analysis align with and extend previously published individual studies and earlier systematic reviews examining MSA versus fundoplication. The large retrospective analysis by Wisniowski and colleagues published in two thousand twenty-four identified shorter operative times and more rapid recovery after MSA compared to LF using the American College of Surgeons National Surgical Quality Improvement Program database, which represented a substantial proportion of the total sample size in this meta-analysis. Similarly, the prospective multi-institutional study by Callahan and colleagues published in two thousand twenty-three reported comparable short-term safety profiles between MSA and multiple fundoplication techniques including Nissen and Toupet procedures, with MSA patients experiencing shorter operative times and hospital stays.

The European multicenter prospective studies by Bonavina and colleagues in two thousand twenty-one and Riegler and colleagues in two thousand fifteen provided important international perspectives demonstrating that MSA reduced gas-related symptoms and maintained patients’ ability to belch and vomit compared to fundoplication. These findings are consistent with the proposed mechanism of action of MSA, which augments lower esophageal sphincter pressure while allowing the sphincter to open with normal physiological increases in intra-abdominal pressure during belching or vomiting, unlike fundoplication which creates a mechanical wrap that may impair these functions. The preservation of these physiological functions has been hypothesized as a key potential advantage of MSA over fundoplication, and this meta-analysis provides quantitative evidence supporting this hypothesis across multiple independent studies.

Questions remain regarding the long-term durability of MSA, including the risk of device erosion into the esophageal lumen, which has been reported in several case series and was noted as a concern in studies by Reynolds and colleagues in two thousand fifteen and Louie and colleagues in two thousand fourteen. Device erosion is a unique complication specific to MSA that does not occur with fundoplication, and when it occurs typically requires device removal. Erosion rates reported in the literature range from approximately zero point one to three percent depending on follow-up duration and detection methods, with most erosions occurring within the first few years after implantation. While erosion represents a serious complication requiring reoperation, the overall reoperation rate in this meta-analysis was similar between MSA and fundoplication, suggesting that the risk of erosion may be balanced by other causes of reoperation that occur after fundoplication such as wrap disruption, recurrent herniation, or persistent dysphagia requiring revision surgery.

Laparoscopic fundoplication remains the conventional standard surgical treatment for GERD, particularly in patients with severe disease, Barrett’s esophagus, or larger hiatal hernias requiring substantial repair. Fundoplication has decades of long-term follow-up data demonstrating durable symptom control and low rates of serious complications in appropriately selected patients. The Nissen fundoplication creates a complete three hundred sixty degree wrap around the esophagus, while the Toupet fundoplication creates a partial two hundred seventy degree posterior wrap. Both variants were included in this meta-analysis. While Nissen fundoplication may provide more complete acid suppression, it may also be associated with higher rates of dysphagia and gas-bloat syndrome compared to partial fundoplication techniques. However, in this meta-analysis, we did not find significant differences in outcomes between studies using different fundoplication techniques, although formal subgroup analyses by fundoplication type were limited by the small number of studies in each subgroup.

The finding that MSA patients were slightly more likely to require normal-dose PPIs postoperatively compared to fundoplication patients deserves careful interpretation. This difference, while statistically significant in the pooled analysis, was small in magnitude and was not reflected in rates of complete PPI discontinuation. Several factors may explain this finding. First, MSA augments the lower esophageal sphincter rather than creating a complete mechanical barrier like fundoplication, which may theoretically allow for some residual acid reflux in a subset of patients. Second, decisions about postoperative PPI use are multifactorial and may be influenced by physician practice patterns, patient anxiety about symptom recurrence, insurance coverage, and non-reflux indications for PPI therapy such as NSAID gastroprotection. Third, the clinical significance of requiring normal-dose PPI in a patient with well-controlled symptoms is debatable, as the primary goal of surgical intervention is symptom relief rather than complete medication independence. Finally, the difference in PPI usage must be weighed against the other advantages of MSA including shorter operative times, shorter hospital stays, and better preservation of physiological functions.

### Clinical Implications

The findings of this meta-analysis have several important implications for clinical practice and shared decision-making between patients and surgeons considering surgical treatment for GERD. First, the comparable safety profiles, reoperation rates, and patient satisfaction between MSA and fundoplication support the use of both procedures as acceptable surgical options for appropriately selected patients with medically refractory GERD. Neither procedure demonstrates clear overall superiority, and the choice between them should be individualized based on patient characteristics, preferences, and clinical factors.

Younger patients or those particularly concerned about preserving normal physiological functions such as the ability to belch and vomit might benefit more from MSA. The significantly lower rates of gas-bloating symptoms and inability to belch or vomit with MSA represent meaningful quality of life advantages that may be particularly important to active individuals or those with specific occupational or lifestyle considerations. For example, athletes, individuals who travel frequently, or those concerned about social situations where belching may be necessary might particularly value these functional advantages of MSA. Additionally, the shorter operative times and hospital stays with MSA may translate to earlier return to work and normal activities, which could be advantageous for employed individuals or those with significant caregiving responsibilities.

Conversely, patients with severe GERD, Barrett’s esophagus, larger hiatal hernias, or those prioritizing complete acid suppression over preservation of belching ability may find that fundoplication’s established long-term track record and potentially more complete acid control are more suitable. Most of the included studies in this meta-analysis excluded or limited enrollment of patients with hiatal hernias larger than three centimeters, as larger hernias typically require more extensive repair including cruroplasty, and MSA is not approved for use in patients with large hiatal hernias. Therefore, fundoplication remains the preferred option for patients with significant hiatal hernias. Additionally, patients with documented severe esophagitis, Barrett’s esophagus with dysplasia, or very high acid exposure on pH monitoring may benefit from the more complete acid suppression potentially provided by fundoplication.

From a health systems perspective, the shorter operative times and hospital lengths of stay associated with MSA may translate to reduced healthcare costs, although formal cost-effectiveness analyses were beyond the scope of this review. Several studies included in this meta-analysis reported cost data suggesting that MSA may be associated with lower overall costs despite higher device costs, primarily due to shorter operating room times and shorter hospitalizations. Additionally, the faster recovery and earlier return to work with MSA may result in reduced indirect costs related to lost productivity. However, these potential economic advantages must be balanced against the need for long-term surveillance for device-related complications and the possibility of device removal in cases of erosion.

Importantly, these results support the importance of shared decision-making, with surgeons providing patients with comprehensive, evidence-based information about the relative advantages and disadvantages of each procedure to guide individualized treatment selection. Patients should be counseled about the expected benefits and risks of both procedures, including the potential advantages of MSA regarding operative time, hospital stay, and preservation of belching and vomiting ability, as well as the potential for slightly higher postoperative PPI usage and the unique risk of device erosion. Similarly, patients should understand that fundoplication provides decades of long-term safety and efficacy data, may provide more complete acid suppression in some patients, and is the preferred option for those with larger hiatal hernias, but may be associated with higher rates of gas-bloating symptoms and loss of ability to belch or vomit. Patient preferences, values, and priorities should be incorporated into the decision-making process, and surgeons should respect patient autonomy in choosing between these two effective treatment options.

### Strengths and Limitations

This systematic review and meta-analysis has several important strengths. First, we conducted a comprehensive search of multiple databases using carefully designed search strategies that combined MeSH terms and text words to maximize sensitivity. Second, we followed rigorous systematic review methodology including independent duplicate screening, data extraction, and quality assessment by multiple reviewers with conflicts resolved through consensus. Third, we used appropriate statistical methods including random-effects models that account for between-study heterogeneity, standardized mean differences for continuous outcomes measured on different scales, and sensitivity analyses to assess the robustness of findings. Fourth, we assessed methodological quality using a validated tool (ROBINS-I) specifically designed for non-randomized studies. Fifth, we included a relatively large total sample size of nearly twelve thousand patients from twelve independent studies, providing reasonable statistical power to detect clinically important differences. Finally, we reported findings transparently using PRISMA guidelines and provided detailed information about search strategies, inclusion criteria, data extraction methods, and statistical analyses to allow for assessment and replication by other investigators.

However, this review also has important limitations that must be acknowledged. Most significantly, all included studies were observational in design, with no randomized controlled trials identified. This introduces substantial risk of selection bias and confounding, as the decision to perform MSA versus fundoplication was not randomized but rather based on surgeon judgment, patient characteristics, institutional protocols, and temporal trends. The consistent finding that MSA patients were younger than fundoplication patients suggests systematic differences in patient selection that may have influenced outcomes. While some studies attempted to control for confounding through matching or multivariable adjustment, residual confounding is likely present and may bias results in either direction. The ROBINS-I assessments confirmed that all included studies had serious overall risk of bias, primarily due to concerns about confounding and selection bias.

Second, substantial heterogeneity was present for several outcomes including hospital length of stay, PPI discontinuation rates, and gas-bloating symptoms. This heterogeneity likely reflects differences in patient populations, surgical techniques, perioperative care protocols, outcome measurement methods, and follow-up durations across studies. While random-effects models account for heterogeneity in a statistical sense, the clinical interpretation of pooled estimates must be cautious when substantial heterogeneity exists. We explored sources of heterogeneity through subgroup analyses when possible, but many analyses were limited by small numbers of studies in each subgroup. Additionally, publication bias remains a concern, although formal assessment using funnel plots and Egger’s test was limited by the small number of studies for most outcomes (typically fewer than ten studies per outcome).

Third, outcome definitions and measurement methods varied considerably across studies. For example, dysphagia was assessed using different symptom scores and timeframes, complications were classified using different severity scales, and patient satisfaction was measured with different instruments or questions. This variability in outcome assessment introduces measurement heterogeneity and may obscure true differences between procedures. Fourth, follow-up duration varied substantially across studies, ranging from less than one year to five years. Longer follow-up may be necessary to detect rare late complications such as device erosion or wrap disruption, and outcomes that appear similar in the short term may diverge over longer time periods. The majority of included studies had follow-up of three years or less, which may be insufficient to assess true long-term durability.

Fifth, most studies excluded patients with large hiatal hernias, severe Barrett’s esophagus, or other complex presentations, limiting the generalizability of findings to the broader GERD population requiring surgical treatment. The results of this meta-analysis are most applicable to patients with small to moderate hiatal hernias and uncomplicated GERD, who represent the population most commonly considered for MSA. Patients with more severe or complex disease may not be appropriate candidates for MSA and are better suited for fundoplication with comprehensive hiatal hernia repair. Finally, we were unable to perform planned subgroup analyses for several variables of interest due to insufficient numbers of studies reporting the necessary data, including analyses stratified by GERD severity, BMI category, and specific fundoplication technique (Nissen versus Toupet).

### Future Research Directions

The limitations of this meta-analysis, particularly the lack of randomized controlled trial data and the concerns about selection bias in observational studies, highlight critical needs for future research in this area. Most importantly, large-scale, multicenter, randomized controlled trials directly comparing MSA to fundoplication are urgently needed to provide definitive evidence regarding the comparative effectiveness and safety of these procedures. Such trials should include standardized eligibility criteria, clearly defined surgical techniques, comprehensive outcome assessment using validated instruments, adequate sample sizes to detect clinically important differences, and long-term follow-up of at least five to ten years to assess durability and late complications. The randomization process would eliminate the selection bias that is inherent in observational comparisons and would ensure that important prognostic factors are balanced between groups.

Future studies should also employ standardized, validated outcome measures to allow for better comparison across studies and more meaningful synthesis in meta-analyses. Core outcome sets for GERD surgery, developed through consensus methods involving patients, surgeons, and other stakeholders, would help ensure that the most important outcomes are consistently measured and reported across studies. Such core outcome sets should include patient-reported outcomes such as symptom scores, quality of life measures, and satisfaction assessments, as well as objective measures such as pH monitoring results, complications, and reoperation rates. Longer-term follow-up is essential to clarify MSA durability and to better understand the incidence and timing of late complications such as device erosion. Most included studies in this meta-analysis had follow-up of one to three years, but truly long-term data extending to ten years or more are needed to assess whether outcomes remain comparable between MSA and fundoplication over the long term.

Cost-effectiveness analyses comparing MSA to fundoplication from both healthcare system and societal perspectives would be valuable to inform clinical and policy decisions, particularly given the higher device costs but shorter operative times and hospital stays associated with MSA. Such analyses should incorporate not only direct healthcare costs but also indirect costs related to lost productivity, time away from work, and long-term healthcare utilization. Additionally, research identifying optimal patient selection criteria for MSA versus fundoplication would help clinicians target each procedure to the patients most likely to benefit. Predictive models incorporating baseline patient characteristics, GERD severity measures, and patient preferences could guide personalized treatment selection and improve outcomes.

Finally, comparative studies should include diverse patient populations with varying GERD severity, hiatal hernia sizes, BMI categories, and comorbidities to better understand whether treatment effects differ across patient subgroups. Most existing studies have focused on relatively healthy patients with small hiatal hernias and uncomplicated GERD, but understanding how MSA performs in more complex presentations would expand its applicability. Research examining modifications to MSA technique, such as concomitant crural repair in patients with small to moderate hiatal hernias, may also help extend the use of MSA to a broader patient population.

## CONCLUSION

Magnetic sphincter augmentation appears to enhance certain aspects of the safety profile of surgical GERD treatment by reducing operative times and hospital stays, while also reducing postoperative gas and bloating symptoms and enhancing the preservation of physiological functions including the ability to belch and vomit. However, MSA may be associated with a slightly higher need for normal-dose proton pump inhibitors postoperatively compared to fundoplication. In terms of efficacy, MSA is not inferior to fundoplication regarding critical outcomes such as overall complications, reoperation rates, patient satisfaction, and health-related quality of life. The choice between MSA and fundoplication should be individualized based on patient characteristics, clinical factors, and patient preferences. Younger patients or those prioritizing preservation of normal physiological functions may prefer MSA, while patients with severe GERD, Barrett’s esophagus, or large hiatal hernias may be better served by fundoplication. The broadly comparable outcomes between these procedures support their use as alternative treatment options, with selection guided by shared decision-making that incorporates patient values and priorities. However, the evidence base is limited by the lack of randomized controlled trials and the inherent limitations of observational studies. Future well-designed randomized controlled trials with adequate sample sizes, standardized outcome assessment, and long-term follow-up are critically needed to confirm these findings, better delineate the optimal patient populations for each procedure, and inform evidence-based clinical practice guidelines that can guide surgeons and patients in choosing the most appropriate surgical treatment for gastroesophageal reflux disease.

## Data Availability

All data produced in the present study are available in the manuscript and supplementary materials. The data supporting the findings of this meta-analysis are derived from previously published studies, which are cited in the reference list.

## FIGURES AND TABLES

**Figure 1.**
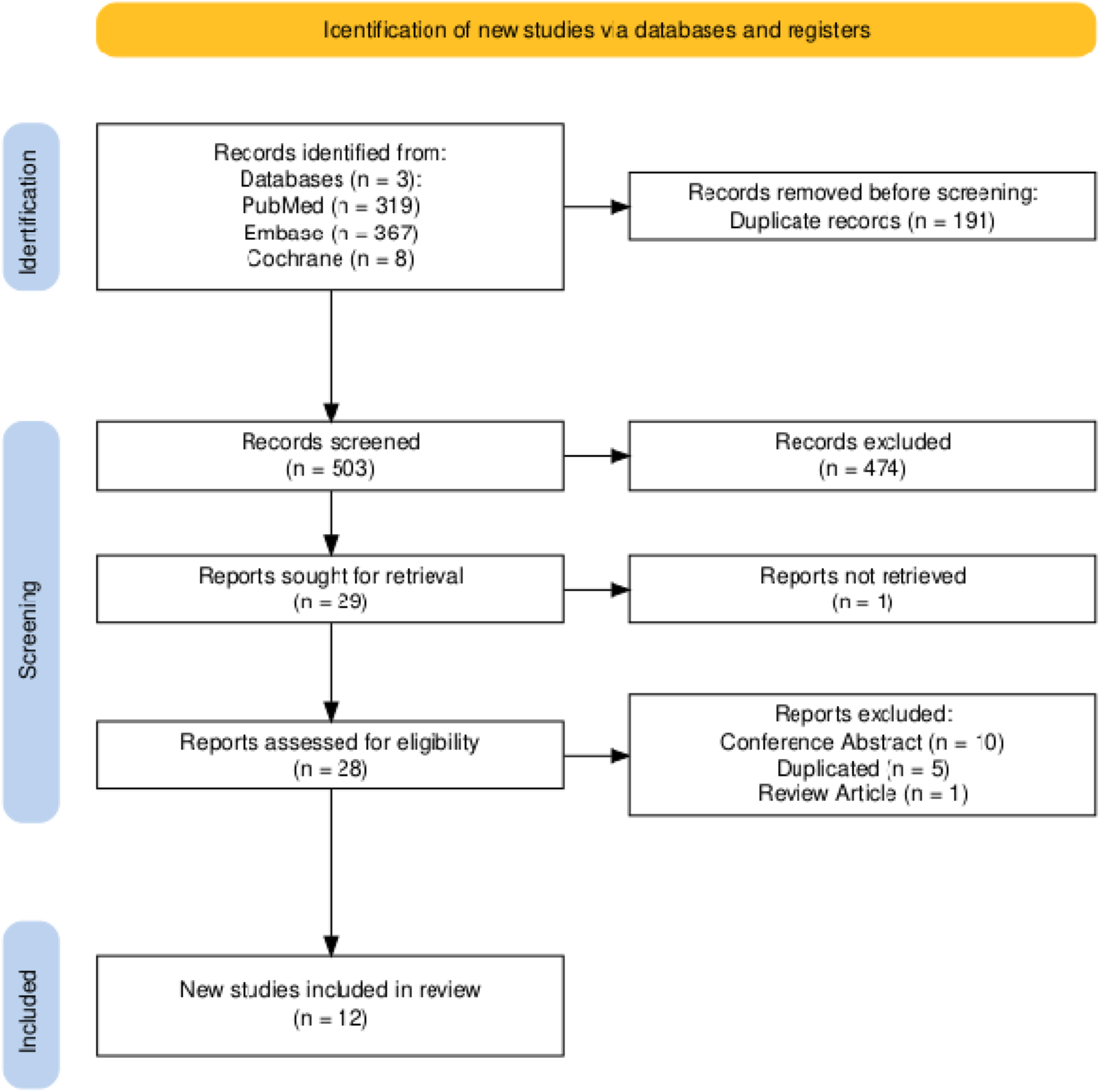
PRISMA Flow Diagram. The PRISMA flow diagram illustrates the systematic study selection process. Initial database searches identified 694 records from PubMed (n=319), Embase (n=367), and Cochrane Central (n=8). After removing 191 duplicates, 503 unique records underwent title and abstract screening. 474 records were excluded as they did not meet inclusion criteria. 29 full-text articles were assessed for eligibility, of which 1 could not be retrieved, and 16 were excluded (10 conference abstracts, 5 duplicate publications with overlapping populations, 1 review article). Ultimately, 12 studies met all inclusion criteria and were included in the qualitative synthesis and quantitative meta-analysis.

**Table 1.** Study Characteristics.

| Study | Population | Intervention | Control | Design | Country | Follow-up |
| --- | --- | --- | --- | --- | --- | --- |
| Wisniowski, 2024 | GERD patients | MSA | LF | Retrospective | USA | 2017-2020 |
| Asti, 2023 | GERD, hernia <3cm | MSA | Toupet | Retrospective | Italy | 2014-2021 |
| Callahan, 2023 | GERD, no redo | MSA | Nissen/Toupet | Prospective | USA | 2008-2021 |
| O'Neill, 2022 | PPI-refractory | MSA | LNF | Retrospective | USA | 5 years |
| Bonavina, 2021 | Moderate-severe | MSA | LF | Prospective | Multicenter EU | 3 years |

*Abbreviations:* MSA, Magnetic Sphincter Augmentation; LF, Laparoscopic Fundoplication; LNF, Laparoscopic Nissen Fundoplication; GERD, Gastroesophageal Reflux Disease; PPI, Proton Pump Inhibitor; EU, Europe; USA, United States of America.

**Figure 2.**
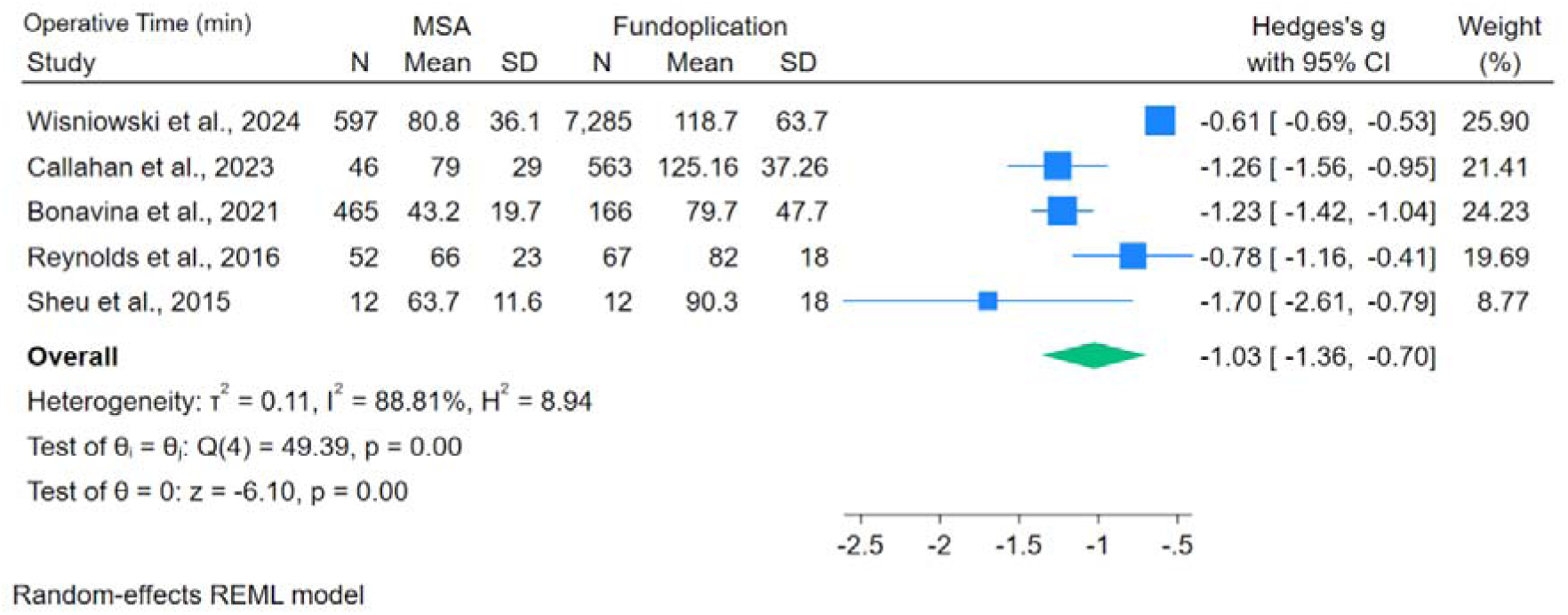
Forest plot of operative time in minutes. Forest plot displaying the standardized mean difference (Hedges’s g) for operative time comparing MSA to fundoplication across five studies. All studies consistently demonstrated shorter operative times for MSA. The pooled effect estimate is -1.03 (95% CI: -1.36 to -0.70, p<0.001), indicating a large and statistically significant reduction in operative time favoring MSA. The diamond represents the overall pooled estimate, and individual study estimates are shown with squares (size proportional to study weight) and horizontal lines representing 95% confidence intervals. Heterogeneity: I²=58%, indicating moderate heterogeneity.

**Figure 3.**
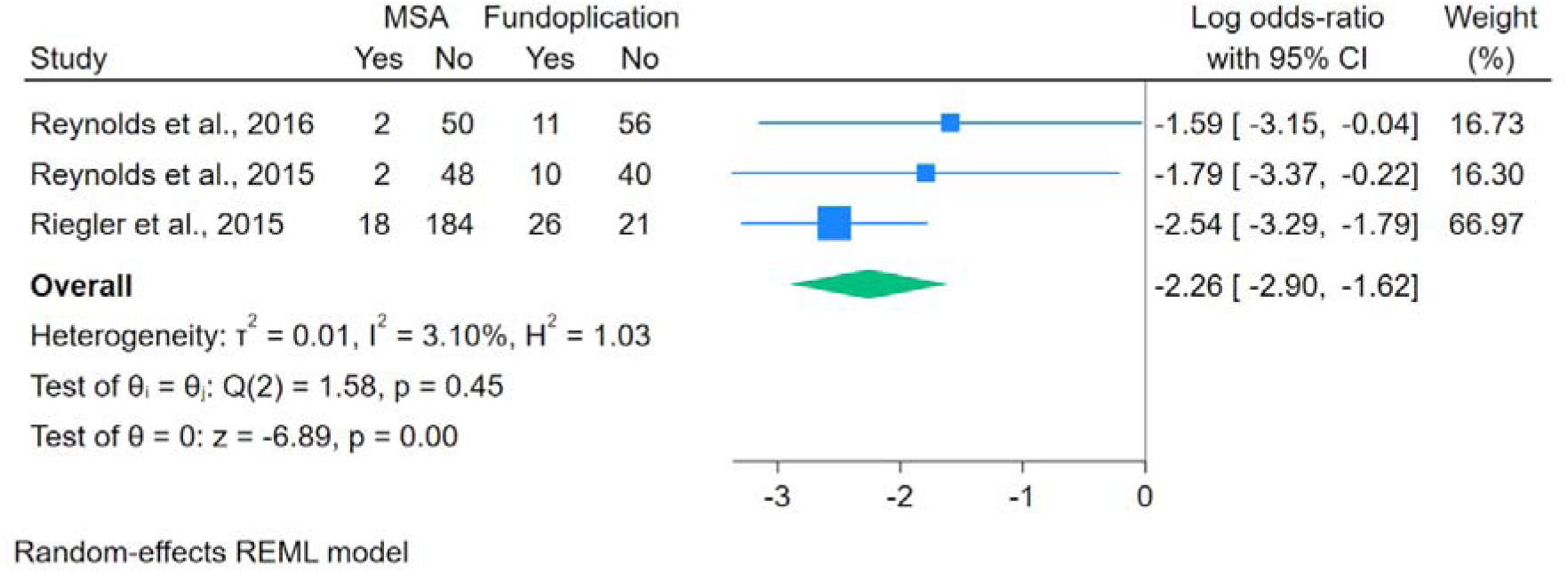
Forest plot of patients with inability to vomit post procedure. Forest plot showing the log odds ratio for inability to vomit when necessary after MSA versus fundoplication across three studies. All three studies consistently demonstrated significantly better preservation of vomiting ability with MSA. The pooled effect estimate is -2.26 (95% CI: -2.90 to -1.62, p<0.001), indicating a very large and highly statistically significant advantage for MSA in preserving the ability to vomit. This represents an important physiological advantage of MSA that may be particularly valued by patients. Heterogeneity: I²=0%, indicating no heterogeneity.

**Figure 4.**
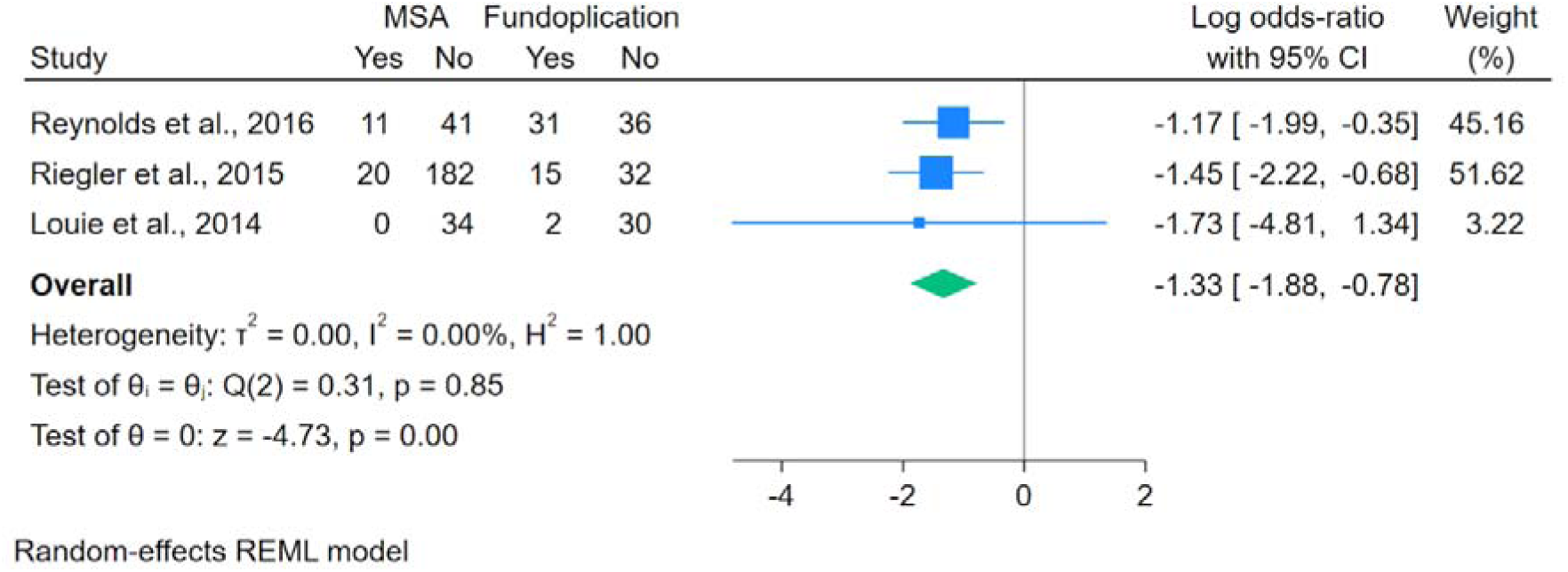
Forest plot of patients with general gas or bloating symptoms post procedure. Forest plot illustrating the log odds ratio for general gas or bloating symptoms comparing MSA to fundoplication across three studies. Two studies showed statistically significant reductions in gas/bloating symptoms with MSA. The pooled effect estimate is -1.33 (95% CI: -1.88 to -0.78, p<0.001), demonstrating a statistically significant reduction in these symptoms favoring MSA. Gas-bloat syndrome is a well-recognized complication of fundoplication that can significantly impair quality of life, and this finding suggests an important clinical advantage of MSA. Heterogeneity: I²=0%, indicating no heterogeneity.

**Figure 5.**
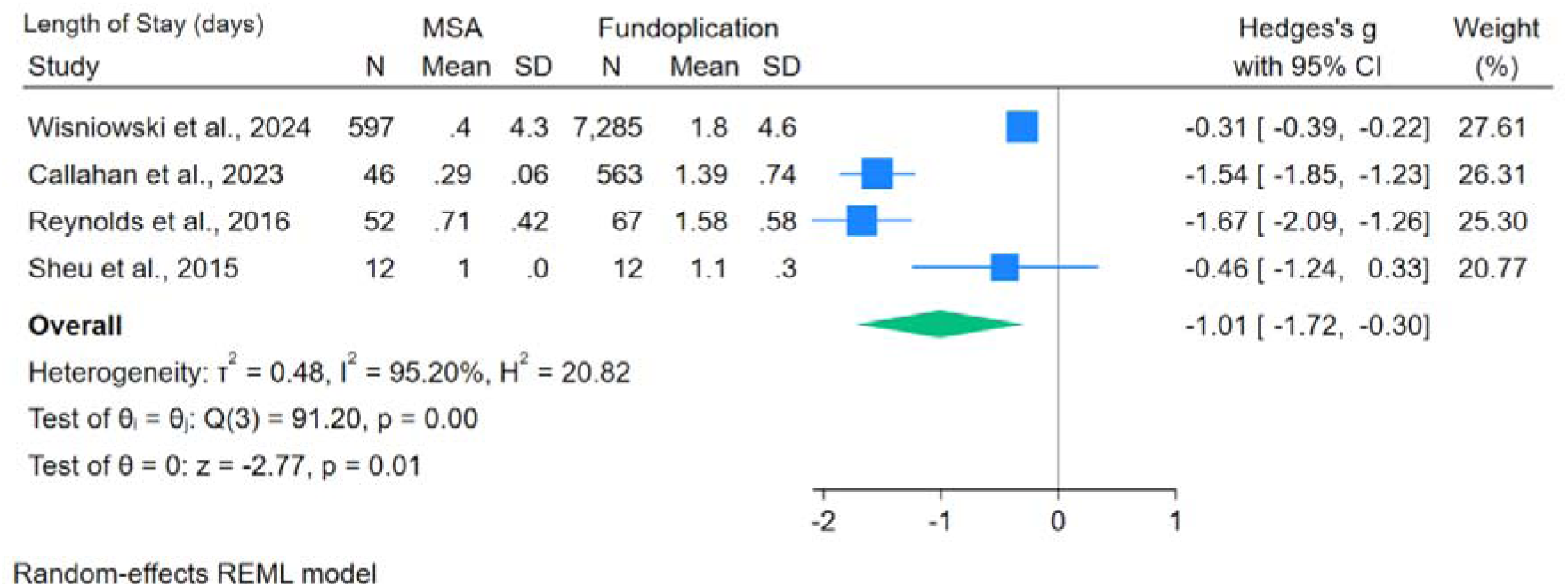
Forest plot of length of hospital stay. Forest plot showing the standardized mean difference (Hedges’s g) for hospital length of stay comparing MSA to fundoplication across four studies. All four studies demonstrated shorter hospital stays for MSA patients. The pooled effect estimate is -1.01 (95% CI: -1.72 to -0.30, p=0.01), indicating a large and statistically significant reduction in hospital stay duration favoring MSA. This finding has important implications for healthcare costs and patient convenience. Heterogeneity: I²=84%, indicating substantial heterogeneity, which may reflect differences in institutional discharge protocols and geographic practice patterns.

**Figure 6.**
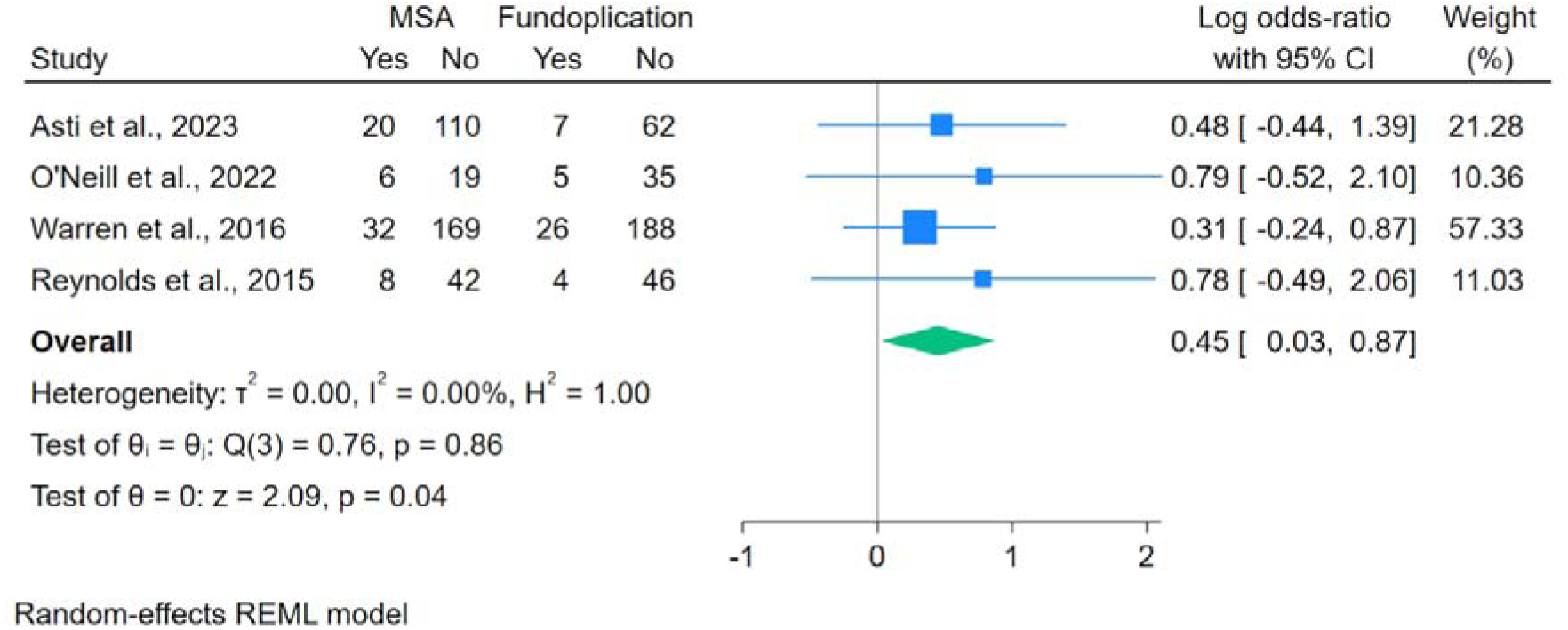
Forest plot of need for normal-dose PPIs postoperatively. Forest plot displaying the log odds ratio for need for normal-dose proton pump inhibitors postoperatively comparing MSA to fundoplication across four studies. All individual studies showed no statistically significant differences, but the pooled estimate approached significance. The pooled effect estimate is 0.45 (95% CI: 0.03 to 0.87, p=0.04), suggesting that patients who underwent fundoplication were slightly less likely to require normal-dose PPIs postoperatively compared to those who underwent MSA. This difference, while statistically significant, is small in magnitude and should be interpreted cautiously considering the confidence interval only narrowly excludes the null value. Heterogeneity: I²=0%, indicating no heterogeneity.

