## Supplemental files for "Comparative Outcomes of Laparoscopic Fundoplication and Magnetic Sphincter Augmentation for GERD: A Systematic Review and Meta-Analysis"

### APPENDIX A: DATA EXTRACTION SHEET

This appendix presents the complete data extraction sheet used for systematic extraction of data from all included studies. Two independent reviewers extracted data using this standardized form, with discrepancies resolved through discussion with a third reviewer. The data extraction form was piloted on three studies before implementation to ensure clarity and completeness of all fields.

#### Section 1: Study Identification and Characteristics

| **Data Field** | **Description and Format** |
| --- | --- |
| Study ID | Unique identifier assigned to each study (e.g., Wisniowski2024, Asti2023) |
| First Author | Last name of first author (text) |
| Publication Year | Year of publication (YYYY format) |
| Journal | Name of journal (text) |
| Study Design | Type of study: Randomized Controlled Trial / Prospective Cohort / Retrospective Cohort / Case-Control / Matched-Pair (dropdown selection) |
| Country | Country or countries where study was conducted (text, use 'Multicenter' for multi-country studies) |
| Institution(s) | Name(s) of participating institution(s) (text) |
| Study Period - Start | Date when patient enrollment began (MM/YYYY format) |
| Study Period - End | Date when patient enrollment ended (MM/YYYY format) |
| Follow-up Duration | Mean or median follow-up duration in months (numeric) and range if reported |
| Funding Source | Source of funding: None / Institutional / Industry / Government / Not Reported (dropdown selection with text field for details) |
| Conflicts of Interest | Reported conflicts: Yes (specify) / No / Not Reported (dropdown with text field) |

#### Section 2: Patient Population and Sample Size

| **Data Field** | **Description and Format** |
| --- | --- |
| Total Sample Size | Total number of patients included in study (numeric) |
| MSA Group Sample Size | Number of patients who received magnetic sphincter augmentation (numeric) |
| Fundoplication Group Sample Size | Number of patients who received laparoscopic fundoplication (numeric) |
| Inclusion Criteria | Study-specific inclusion criteria as reported by authors (text) |
| Exclusion Criteria | Study-specific exclusion criteria as reported by authors (text) |
| GERD Diagnosis Criteria | Method(s) used to diagnose GERD: Clinical symptoms / Endoscopy / pH Monitoring / Manometry (multiple selections possible with text field for details) |

#### Section 3: Baseline Demographics (MSA Group)

| **Data Field** | **Description and Format** |
| --- | --- |
| Age MSA - Mean | Mean age in years (numeric with decimal) |
| Age MSA - Standard Deviation | Standard deviation of age (numeric with decimal) |
| Age MSA - Median | Median age if reported instead of mean (numeric) |
| Age MSA - IQR or Range | Interquartile range or full range (text format: 'Q1-Q3' or 'min-max') |
| Female Patients MSA - Number | Number of female patients (numeric) |
| Female Patients MSA - Percentage | Percentage of female patients (numeric with decimal) |
| Male Patients MSA - Number | Number of male patients (numeric) |
| Male Patients MSA - Percentage | Percentage of male patients (numeric with decimal) |
| BMI MSA - Mean | Mean body mass index in kg/m² (numeric with decimal) |
| BMI MSA - Standard Deviation | Standard deviation of BMI (numeric with decimal) |

**Note:** Section 3 is repeated identically for the Fundoplication Group with field names changed from 'MSA' to 'Fundoplication'. All demographic variables were collected for both treatment groups to allow for baseline comparison.

#### Section 4: Baseline Clinical Characteristics (Both Groups)

| **Data Field** | **Description and Format** |
| --- | --- |
| GERD-HRQL Score - Mean | Mean GERD Health-Related Quality of Life score at baseline (numeric, scale 0-50, extracted separately for MSA and Fundoplication groups) |
| GERD-HRQL Score - SD | Standard deviation of GERD-HRQL score (numeric with decimal) |
| DeMeester Score - Mean | Mean DeMeester composite score from 24-hour pH monitoring (numeric, normal <14.7, extracted separately for both groups) |
| DeMeester Score - SD | Standard deviation of DeMeester score (numeric with decimal) |
| Percent Time pH<4 - Mean | Mean percentage of time with esophageal pH less than 4 on 24-hour monitoring (numeric, normal <4%, extracted separately for both groups) |
| Percent Time pH<4 - SD | Standard deviation of percent time pH<4 (numeric with decimal) |
| LES Pressure - Mean | Mean lower esophageal sphincter pressure in mmHg from esophageal manometry (numeric, normal 10-30 mmHg, extracted separately for both groups) |
| PPI Duration - Mean (days) | Mean duration of PPI therapy before surgery in days (numeric, extracted separately for both groups) |
| PPI Duration - SD | Standard deviation of PPI duration (numeric) |
| Hiatal Hernia Present - Number | Number of patients with hiatal hernia present on endoscopy or imaging (numeric, extracted separately for both groups) |
| Hiatal Hernia Size - Mean (cm) | Mean size of hiatal hernia in centimeters (numeric with decimal, extracted separately for both groups) |
| Hiatal Hernia Size - SD | Standard deviation of hiatal hernia size (numeric with decimal) |
| Esophagitis Present - Number | Number of patients with esophagitis on endoscopy (numeric, extracted separately for both groups) |
| Esophagitis Grade | LA Classification if reported: Grade A / B / C / D (text with number of patients in each grade) |
| Barrett's Esophagus - Number | Number of patients with Barrett's esophagus (numeric, extracted separately for both groups) |

**Note:** All fields in Section 4 were extracted separately for MSA and Fundoplication groups to enable baseline comparison. Additional clinical characteristics such as previous abdominal surgeries, comorbidities (diabetes, hypertension, etc.), smoking status, and alcohol use were also extracted when reported.

Sections 5 through 10 of the data extraction sheet include detailed fields for Surgical Characteristics, Intraoperative Data, Postoperative Outcomes (Complications, Symptoms, Quality of Life, PPI Usage), Reoperation Data, Patient Satisfaction Measures, and Long-term Follow-up Outcomes. Each section contains 15-25 specific data fields with defined formats and units. The complete data extraction sheet spans over 200 individual data fields to ensure comprehensive and systematic extraction of all relevant information from included studies.
